# Accurate Non-invasive Mass and Temperature Quantifications with Spectral CT

**DOI:** 10.1101/2022.02.18.22271054

**Authors:** Leening P. Liu, Matthew Hwang, Matthew Hung, Michael C. Soulen, Thomas P. Schaer, Nadav Shapira, Peter B. Noël

## Abstract

Spectral CT has been increasingly implemented clinically for its better characterization and quantification of materials through its multi-energy results. It also facilitates calculation of physical density utilizing the Alvarez-Macovski model without approximations. These spectral physical density quantifications allow for non-invasive mass measurements and temperature evaluations by manipulating the definition of physical density and thermal volumetric expansion, respectively. To develop the model, original and parametrized versions of the Alvarez-Macovski model and electron density-physical density model were validated with a phantom. The best physical density model was then implemented on clinical spectral CT scans of *ex vivo* bovine muscle to determine the accuracy and effect of acquisition parameters on mass measurements. In addition, the relationship between physical density and changes in temperature was evaluated by scanning and subjecting the tissue to a range of temperatures. A linear fit utilizing the thermal volumetric expansion was performed to assess the correlation. The parametrized Alvarez-Macovski model performed best in both model development and validation with errors within ±0.02 g/mL. As observed with muscle, physical density was not significantly affected by dose and acquisition mode but was slightly affected by collimation. These effects were also reflected in mass measurements, which demonstrated accuracy with a maximum percent error of 0.34%, further validating the physical density model. Furthermore, physical density was strongly correlated (R of 0.9781) to temperature changes through thermal volumetric expansion. Accurate and precise spectral physical density quantifications enable non-invasive mass measurements for pathological detection and temperature evaluation for thermal therapy monitoring in interventional oncology.

## I. Introduction

**W**ITHits improved image quality, tissue characterization, and material quantification capabilities, spectral computed tomography (CT) has been increasingly adopted and implemented in the clinical setting. It provides spectral results, including virtual mono-energetic images (VMI), iodine density maps, effective atomic number (Zeff) maps, and electron density (ED) maps, that have enabled quantitative evaluation of clinical scenarios [1], [2]. They have been critical for a growing number of applications [3]–[6]. For example, improved lesion characterization requires separation of potential malignant tumors from benign lesions and cysts to determine the necessary clinical care [7]. By examining relevant spectral results, specifically iodine density maps and VMI of different energies for lesion characterization [8], [9], confidence and accuracy in quantification and material composition increase, directly leading to improved diagnostics [10], [11]. Overall, these spectral quantifications not only supplement images with more consistent and quantitative metrics for diagnostic imaging but also enable utilization of different spectral results to calculate relevant quantities, such as physical density. Both advantages may also expand the use of spectral results to interventional procedures.

Physical density quantifications have been generated with spectral CT to describe the relationship between a material’s mass and volume in non-clinical applications, particularly petroleum and mineral analysis [12]. While a linear relationship has been used to characterize the relationship between Hounsfield Units (HU) and physical density with conventional CT [13]–[15], spectral CT has allowed the development of more elaborate models by acquiring attenuation from two or more different polychromatic spectra [16]. One such model stems from the Alvarez-Macovski (AM-PD) model that describes material-dependent attenuation at a given energy [16]. It is expressed as a linear combination of the two main physical effects responsible for x-ray attenuation (photoelectric effect and Compton scattering), both of which are a function of physical density, atomic number, and atomic mass. Previous implementations of this model for physical density quantifications handled high and low energy attenuation maps from spectral CT as inputs into the model [12], [17]. Because of the polychromatic spectra used to generate high and low energy attenuation maps, the AM-PD model requires approximations and/or information about the spectra to accommodate the model’s use of attenuation at a single energy. An alternative that has not been investigated before with AM-PD is taking advantage of clinically available quantitative spectral results to serve as input into the model. This approach removes the need for simplifications to the AM-PD model as attenuation at a single energy and atomic number can be substituted by VMI and Zeff maps that are available on all spectral CT platforms [2], [18].

Compared to non-clinical applications, clinical applications of non-invasive physical density maps remain largely unexplored. There are many potential uses for such maps in abdominal, pulmonary, and breast imaging. Many of these applications are associated with diagnostic imaging that would benefit from direct physical density measurements of lesions for material characterization. Physical density maps also enable non-invasive measurements of mass, an indicator of pathology [19], [20]. This measurement can be achieved by summing physical density maps over a segmented volume and multiplying by voxel volume. These metrics differ from the traditionally HU in CT as they not only are physical and intuitive units that can be easily interpreted, but also correspond to physical properties that can be measured by other techniques, allowing for easy comparison. With the ability of spectral CT to measure physical density and weight, both can be become widely use in diagnostic imaging for determining and characterizing possible pathology.

In addition to the relationship between physical density and mass, physical density also reflects changes in temperature, facilitating real-time non-invasive thermometry for monitoring interventional thermal therapy procedures. The relationship between physical density and change in temperature can be described by the principle of thermal volumetric expansion. Thermal volumetric expansion describes expansion in volume, and consequently decreases in physical density with increases in temperature [21]. By exploiting this relationship, physical density quantifications, unlike HU in conventional CT [21], can distinguish temperature and composition changes to enable real-time non-invasive temperature monitoring. This is of high importance for the field of thermal therapy, particularly thermal ablation therapy where successful ablations rely on tumor tissue and a surrounding margin of healthy tissue reaching a lethal threshold of 60 °C [22]. However, in current clinical practice, ablations are performed by interventional radiologists under CT guidance without the ability to monitor temperature. Thus, establishing the relationship between physical density and temperature changes will allow for real-time non-invasive temperature monitoring for interventional radiologist during ablative procedures, resulting in lower recurrence rates and less damage to surrounding tissue.

This study aimed to develop a physical density model utilizing clinically available spectral results as direct inputs into the model without approximations and investigate two clinical applications of spectral physical density quantifications: non-invasive mass measurements and real-time non-invasive temperature monitoring. In addition, non-invasive mass measurements were evaluated for accuracy and the effect of acquisition parameters while non-invasive temperature monitoring was assessed to establish the relationship between temperature and physical density.

## II. Methods

### A. Physical density model development

To develop an accurate model for spectral physical density quantifications, two main models were explored. The first was the AM-PD model that describes energy- and material-dependent attenuation as a linear combination of the photoelectric effect and Compton scattering, as given by

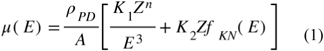

where *E* is photon energy, *ρ*_*PD*_ is physical density, *Z* is atomic number, *A* is atomic mass, *f*_*KN*_(*E*) is Klein-Nishina function, and *K*_1_, *K*_2_ are two material-specific constants [16]. This relation encompasses the two main effects associated with attenuation and does not include Raleigh scattering, which is considered negligent in magnitude and follows a similar energy dependence as described by the photoelectric effect [23]. The other characterizes the relationship between electron density (ED) and physical density (ED-PD model) as given by

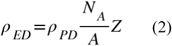

where *N*_*A*_ is Avogadro’s number, ρ_PD_ is physical density, *Z* is atomic number, and is atomic mass. As both models referred to single-element materials, each model required adaption to materials comprised of a mixture of elements by replacing the atomic number and atomic mass with Zeff and effective atomic mass (Aeff). Zeff was represented as a weighted sum of the atomic number as given by

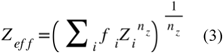

where *Z*_*i*_is the atomic number for the element, *f*_i_ is the fraction of electrons corresponding to the element, and *n*_z_ is the exponent of Zeff. A *n*_z_ of 2.94 was utilized. Aeff, on the other hand, was estimated with a third order polynomial fit between atomic masses and numbers of the first 30 elements. This approximation of the effective atomic number resulted in a R^2^ of 0.9935 and an absolute maximum error of 5.8%.

While both the original relations are widely accepted and utilized, parametrized versions of the AM-PD model and ED-PD model were evaluated in addition to original models to improve the model by accounting for any model assumptions and multi-elemental materials.

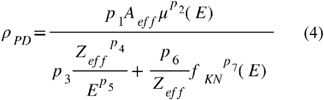

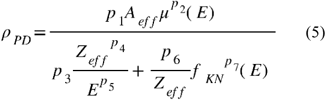

First, to determine the parameters of each parametrized model, anthropomorphic tissues defined by the International Commission on Radiation Units and Measurements (ICRU) Report 44 were described by elemental composition (H, C, N, O, Na, Mg, P, S, Cl, K, Ca, Fe, I), electron density, and physical density [24]. Using this information, attenuation at 70 keV and effective atomic number was calculated using well-accepted material properties, i.e. National Institute of Standard and Technology (NIST) XCOM, and theoretical relations (Eq. 3). Both these quantities in addition to electron density served as input into models, while reported physical density was utilized as ground truth. The calculations were repeated for additional tissues that consisted of varying amounts of iodine (2, 4, 6, 8, 10 mg/mL) added to select tissues (blood, brain, heart, kidney, liver, lung, lymph, muscle, pancreas, urinary bladder) to represent tissues exposed to iodinated contrast media that is typically used in at least 50% of CT examinations [25]. With a total of 180 materials, a least squares fit was applied to the parametrized AM-PD and ED-PD models to determine best fit parameters. Then, physical density for all materials was calculated using all four models, and root mean square error (RMSE) was assessed to characterize model performance.

Physical density models were further validated by applying models to image data of a phantom with known physical density values. This validation utilized a 33 cm diameter tissue characterization phantom (Gammex Model 467, Sun Nuclear, Melbourne, FL, USA) that consisted of 14 tissue-mimicking inserts. These inserts primarily had nominal densities between 0.948 to 1.33 g/mL with tolerances of ± 0.02 as declared by phantom manufacturer. The phantom also included two additional lung inserts with densities of 0.29 and 0.475 g/mL.

The phantom was scanned with a dual-layer spectral detector dual energy CT (IQon Spectral CT, Philips Healthcare, Amsterdam, Netherlands) at two different tube voltages (120, 140 kVp) and two different radiation dose levels (10.6, 34.9 mGy for 120 kVp, 10.3, 31.9 mGy for 140 kVp). Other acquisition parameters are detailed in Table 1. For each scan, clinical spectral results, specifically 70 keV VMIs, electron density maps, and effective atomic number maps, were generated. Using these spectral results, physical density maps were calculated using the original (Eq. 2-3) and parameterized (Eq. 4-5) models, and regions of interest (ROI) with a diameter of 21 mm were placed on each material insert to measure its respective physical density. Values were reported as mean ± standard deviation and illustrated as calculated physical density and as different between calculated and manufacturer-reported nominal density. RMSE was also calculated to determine the performance of the model in validation. The physical density model with the lowest RMSE for physical density from both ICRU44 tissues and the tissue characterization phantom was considered the superior model and utilized for investigating non-invasive mass measurements and temperature evaluation.

**TABLE 1.** Acquisition Parameters for Model Validation, Mass Measurements, and Temperature Evaluation

|  | Model validation | Mass measurement | Temperature evaluation |
| --- | --- | --- | --- |
| Tube voltage | 120/140 | 120 | 120 |
| Exposure time [s] |  | 0.75 |  |
| Pitch | axial | 0.931, axial | axial |
| CTDI<sub>vol</sub> [mGy] | 10.6, 34.9 / 10.3, 31.9 | 15.3, 30.3, 45.5 | 15.2, 30.3, 45.5 |
| Collimation width [mm] | 32x0.625 | 16x0.625, 64x0.625 | 16x0.625 |
| Slice thickness [mm] |  | 2.5 |  |
| Convolution kernel |  | B |  |
| Field of view [mm] |  | 350 |  |
| Matrix size |  | 512x512 |  |
| Pixel spacing [mm] |  | 0.6836x0.6836 |  |

### B. Non-invasive mass measurements

For the first application of spectral physical density quantifications, non-invasive mass measurements of *ex vivo* soft tissue were investigated for feasibility, accuracy, and the effect of scanning parameters on the resulting mass measurements. To accomplish this, *ex vivo* bovine muscle was placed on polyfoam inside the 20 cm bore of a 30×40 cm^2^ phantom (Multi-Energy CT Phantom, Sun Nuclear, Melbourne, FL, USA) and scanned with a dual-layer spectral detector dual energy CT at 120 kVp (Fig. 1A). Scan parameters were varied to examine the impact of different collimations (16×0.625, 64×0.625 mm), radiation dose levels (15.2, 30.3, 45.5 mGy), and CT acquisition modes (axial, helical). No helical scans were acquired with the narrow collimation of 16×0.625 mm at 45.5 mGy due to tube output limitations. Each set of scans was repeated three times. Other acquisition parameters are listed in Table 1. VMI 70 keV and Zeff maps were extracted and inputted into the superior physical density model (parametrized AM-PD model) to acquire physical density maps. Additionally, the muscle was weighed with a precision balance (Fisher Scientific Education Precision Balance, Fisher Scientific, Hampton, NH, USA) before and after scanning to serve as the ground truth mass values.

**Fig. 1.**
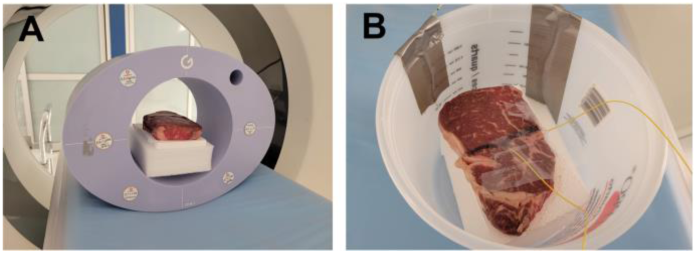
Experimental set up for two potential applications of physical density. An ex vivo bovine muscle was scanned in a multi-energy CT phantom on polyfoam with a dual-layer spectral detector dual energy CT to investigate non-invasive weight measurements (A). To evaluate the changes in physical density in respect to changes in temperature, optical fibers were inserted into muscle as it was subjected to a range of temperatures (B). Scanning occurred simultaneously to temperature measurements and temperature changes.

With physical density maps calculated from the generated/reconstructed spectral results, both physical density values and non-invasive mass measurements were evaluated to characterize the effect of scanning parameters. To assess the effect of scanning parameters on physical density, ROIs with a diameter of 13.6 mm were placed in the center of the muscle on four consecutive slices. Values were reported as mean ± standard deviation over 3 scans (total of 12 slices). A multiple linear regression was fit to determine the effect of collimation, radiation dose, and acquisition mode such that a p-value of 0.05 was considered significant. Non-invasive mass measurements, on the other hand, first required isolation of voxels associated with muscle. A range of thresholds from -950 HU to 100 HU were applied to VMI 70 keV from high dose (45.5 mGy) helical scans. The number of included pixels after thresholding were recorded for each threshold value, and the optimal threshold value was selected such that the increase in threshold no longer resulted in significant reduction in the number of pixels (elbow method). This value (−890 HU), representing the threshold between air and soft tissue, was then employed on VMI 70 keV to generate a mask of pixels associated with muscle that was subsequently applied to corresponding physical density maps from the same scan. Physical density values from included voxels were summed and multiplied by voxel size (0.68 × 0.68 × 2.5 mm^3^) to calculate the total mass of the muscle. Calculated mass was then compared to the average of the two weight measurements to determine its accuracy with different scanning parameters.

### C. Non-invasive temperature evaluations

The second application of spectral physical density quantifications focused on examining the relationship between physical density changes and changes in temperature and its proximity to thermal volumetric expansion. Using the same *ex vivo* muscle utilized for non-invasive mass measurements, optical fibers containing FBG temperature sensors (Fiber Optic Thermometer, Omega Engineering, Norwalk, CT, USA) were inserted into the tissue using 12G medical trocars, which enabled continuous recording of internal temperatures during the experiment (Fig. 1B). The muscle was then positioned in a plastic container. To subject it to a range of temperature, hot water was first poured in to completely submerge the muscle. Then once the muscle reached an equilibrium temperature, ice was added to cool the muscle. During heating and cooling of the sample, it was scanned approximately every minute with a dual-layer spectral detector dual energy CT at a tube voltage of 120 kVp, collimation of 16×0.625 mm, revolution time of 0.75s, and three different dose levels (15.2, 30.3, 45.5 mGy). Other related parameters are listed in Table 1. For each scan, VMI 70 keV and effective atomic number maps were generated and utilized to calculate physical density maps.

To acquire physical density measurements corresponding to recorded temperature values, the location of optical fiber temperature probes was determined by thresholding VMI 70 keV at 90 HU. ROIs with a diameter of 4.1 mm were then automatically placed adjacent to the tip of the optical fiber to measure physical density. In order to relate physical density changes to changes in temperature, the thermal volumetric expansion equation was rearranged and utilized.

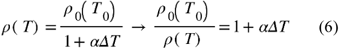

Accordingly, physical density was normalized by dividing the final temporal physical density value (lowest temperature of 22 °C) with the measured physical density at each timepoint. Similarly, the change in temperature was determined relative to the temperature during the final temporal scan. Normalized physical density values were then plotted against change in temperature to illustrate thermal volumetric expansion. A linear regression was fit to the data, where the slope represented the thermal volumetric expansion coefficient. Pearson’s correlation coefficient was determined to characterize the correlation between normalized physical density and change in temperature.

## III. Results

### A. Physical density model development

Between the four physical density models (original AM-PD, parametrized AM-PD, original ED-PD, parametrized ED-PD), the parametrized AM-PD model performed best in model development. In comparison to the original models where errors in physical density for tissues described by ICRU44 were greater than 0.1 g/mL, errors from both the parametrized AM-PD and parametrized ED-PD models were reduced to less than 0.01 g/mL (Fig. 2). Across all tissues, the parametrized AM-PD model outperformed parametrized ED-PD model with a RMSE of 0.007 g/mL compared to parametrized ED-PD’s 0.012 g/mL. Similarly, validation with the tissue characterization phantom (Fig. 3) exhibited that the parametrized AM-PD model performed better than the parametrized ED-PD model. For a majority of the inserts, the parametrized ED-PD model underestimated physical density errors ranging from -0.055 to - 0.025 g/mL. These errors were outside the manufacturer reported tolerance of 0.020 g/mL. The parametrized AM-PD model, on the other hand, demonstrated errors within the reported tolerance, ranging from -0.021 to 0.015 g/mL for all inserts except the two lung inserts (Fig. 3D). Physical density values from the parametrized AM-PD model also did not vary with either tube voltage or dose. With the best performance in both model development and validation, the parametrized AM-PD model was selected as the physical density model to be utilized in following/consecutive experiments.

**Fig. 2.**
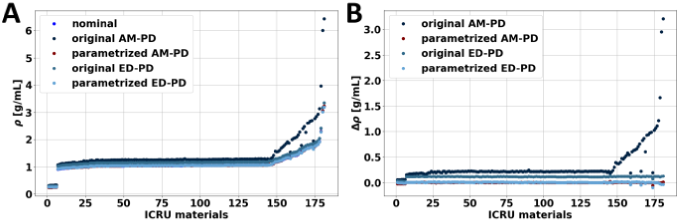
Physical density of ICRU materials calculated with theoretical and parameterized Alvarez-Macovski and ED-PD models. Model development involved materials with a range of physical density values (A). Parametrized AM model performed well in comparison to theoretical questions and parametrized ED-PD model as evidenced by low differences between calculated and nominal values (B).

**Fig. 3.**
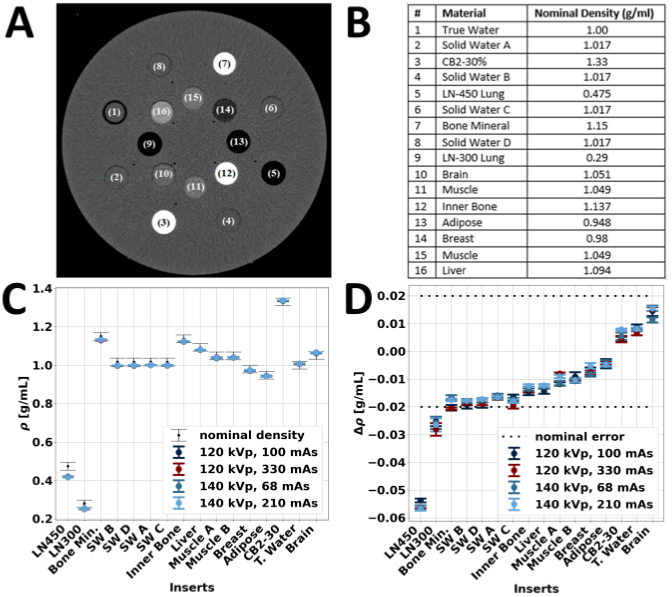
Parametrized AM model validation with DECT measurements of tissue mimicking inserts. A tissue characterization phantom (A) was scanned with multiple tissue mimicking inserts with a range of nominal density values (B). DECT measurements of VMI 70 keV and Zeff were acquired at two different kVps and doses to examine the effect of scan parameters on physical density quantifications (C). Except for the two lung inserts (LN450 and LN300), physical density values were within the nominal error of ± 0.02 g/ml (D, black dashed lines) and did not vary with different scan parameters.

### B. Non-invasive mass measurements

As first observed in the validation of the physical density models and further exemplified in *ex vivo* soft tissue, spectral physical density quantifications were not markedly affected by acquisition parameters (Fig. 4). The effect of radiation dose (p-value of 0.572 and 0.246) and CT acquisition mode (p-value 0.509) were not significant in comparison to the effect of collimation, which resulted in a significant difference (p-value less than 0.000) of 0.003 g/mL between physical density values acquired with a collimation of 16×0.625 mm and 64×0.625 mm. However, the difference of 0.003 g/mL is small, suggesting a minimal effect of collimation on spectral physical density quantifications. The lack of effect of dose and collimation on spectral physical density quantifications were likewise reflected in mass measurements calculated from physical density maps.

**Fig. 4.**
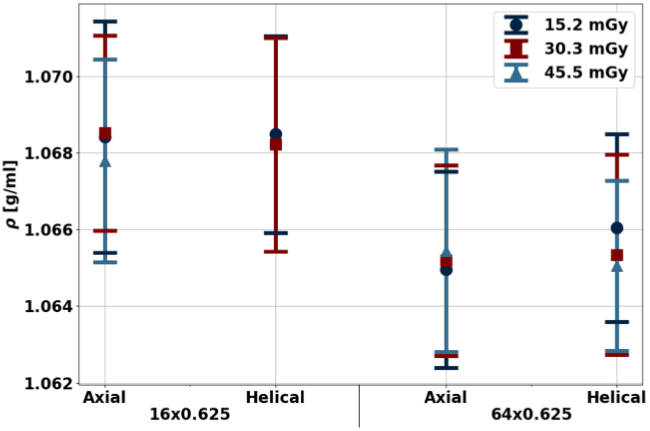
Spectral physical density quantifications at different doses, collimations, and axial/helical scans. With dose matched scans and different collimations, physical density decreased approximately 0.003 g/mL with increased collimation. There was no effect of dose and axial/helical scans on physical density quantifications.

Physical density maps of *ex vivo* soft tissue enabled accurate non-invasive mass measurement independent of dose and collimation (Fig. 5). Between the five different combinations of collimation and dose, measured mass values were within ± 1.1 g of ground truth weight from a precision scale. These differences in mass translated into percent errors of -0.34% and 0.04% for mass measured from scans with a collimation of 16×0.625 and 64×0.625 mm, respectively. The accuracy of non-invasive mass, measurements regardless of acquisition parameters, further validates the physical density model and demonstrates its utility for clinical applications, such as determining the presence of pathology.

**Fig. 5.**
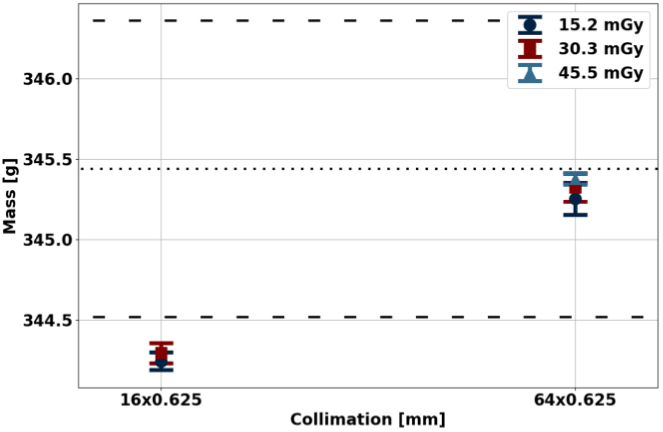
Mass estimation from spectral physical density maps. Estimated mass values were in excellent agreement with measured mass. The dotted line indicates the average mass from two weight measurements (before and after CT scans) while the dashed line represents each independent weight measurement.

### C. Non-invasive temperature evaluation

In addition to non-invasive mass measurements, spectral physical density quantifications enabled an experimental demonstration for non-invasive temperature assessment by exploiting the relationship between physical density and changes in temperature (Fig. 6). Specifically, the linear fit of the manipulated thermal volumetric expansion equation (Eq. 6) demonstrated a slope of 0.00042 ± 0.00001 °C^-1^ and an intercept of 1.0000 ± 0.0003 for temperatures between 22 and 45.5 °C, corresponding to temperature changes of up to 33.5 C (Fig. 7). These fit parameters corresponded to an approximate 0.42% decrease in physical density with an increase in 10°C. A Pearson’s correlation coefficient of 0.9781 demonstrated high linear correlation between normalized physical density and change in temperature, recapitulating thermal volumetric expansion.

**Fig. 6.**
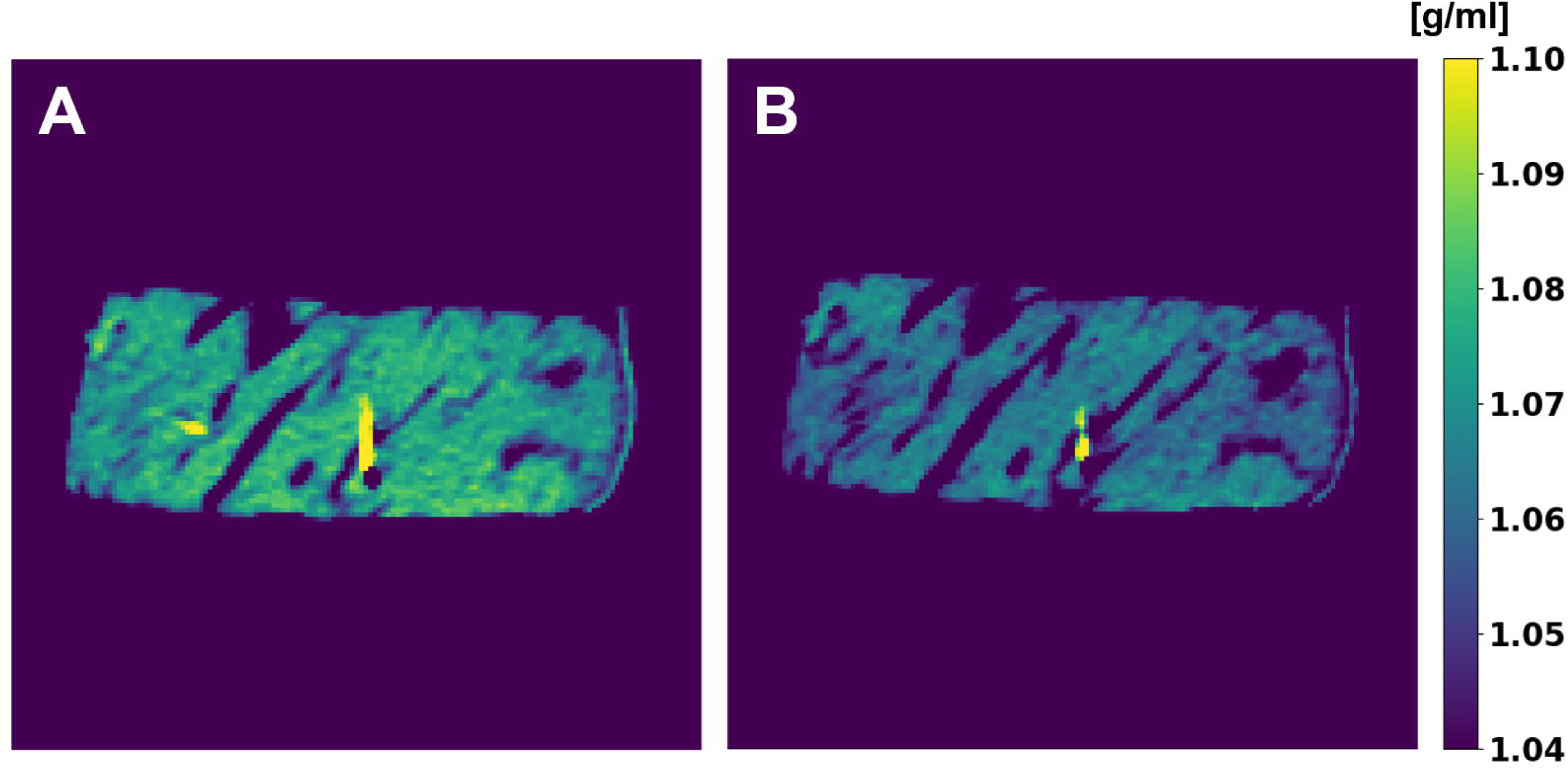
Physical density maps of ex vivo bovine muscle in water (ρ = 1 g/cm^3^) at the lowest (22 °C, A) and highest (45.5 °C, B) temperatures. Increase in temperature resulted in a decrease in physical density. Areas of high constant physical density between temperature yellow) correspond to optical fiber temperature probes placed to obtain ground truth temperature measurements.

**Fig. 7.**
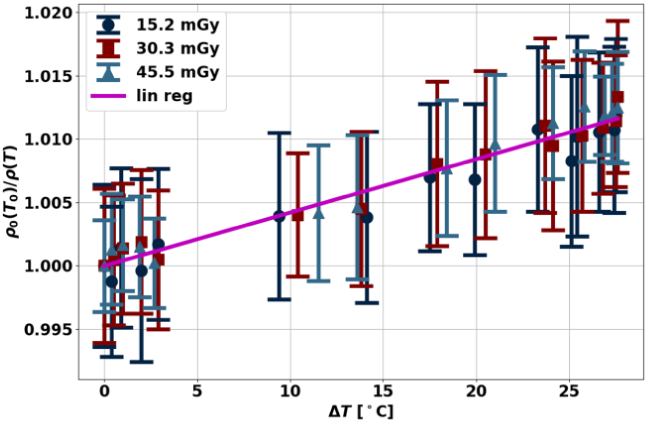
Normalized physical density changes with heating and cooling of ex vivo bovine muscle. The linear relationship between normalized physical density and changes in temperature reflected thermal volumetric expansion, which describes volumetric changes with temperature changes.

## IV. Discussion

Spectral physical density quantifications facilitated accurate non-invasive mass measurements and temperature assessment. These applications were achieved from spectral quantifications derived from the parametrized AM-PD model that were not only accurate for tissue characterization rods and *ex vivo* soft tissue, but also stable with radiation dose, collimation, and CT acquisition mode. Consistent physical density maps translated into measured mass values within ±1.1 g of ground truth weight, demonstrating the accuracy of non-invasive mass measurements. Furthermore, spectral physical density quantifications exemplified a strong linear relationship with temperature changes associated with thermal volumetric expansion, allowing for future non-invasive temperature monitoring capabilities. Overall, both mass measurements and temperature monitoring exhibited only two of many potential clinical applications of spectral physical density quantifications in diagnostic and interventional imaging.

Compared to previous implementations of physical density models, full implementation of the AM-PD model with spectral results did not require approximations and resulted in high accuracy. The model utilized VMI 70 keV, which are clinically available on all spectral CT platform, rather high and low dual energy CT, thereby replacing the need for scanner-specific spectra information or a simplification present in previous iterations of physical density models [12], [15], [17]. This lack of assumptions benefited the accuracy of the model in addition to the strong model foundation (AM-PD model) and parametrization. In particular, the parametrized AM-PD model parameters closely matched the expected values from the original model, recapitulating the original model with minor changes but with improved performance. The increased accuracy with the parametrized AM-PD model likely can be attributed to the characterization of attenuation as the result of only the two main physical effects, the photoelectric effect and Compton scattering. In reality, attenuation also incorporates effects from Raleigh scattering though its contribution is relatively small and follows a similar energy dependence to the photoelectric effect [26]. As a result, parametrization re-encompasses the effect of Rayleigh scattering by slightly altering the original model parameters, thus improving the parametrized AM-PD model’s performance relative to the original model. The high performance of the model not only manifests in the generated physical density maps but also metrics calculated from these maps.

Accurate non-invasive mass measurements highlighted a potential application for pathological identification. Historically, organ weight has not been a clinically utilized parameter as it could not be measured non-invasively. Conversely, autopsies and forensic examinations have regularly evaluated organ weight to determine the presence of pathology [19], [20]. While some pathologies result in distinct changes in size, appearance, and texture with gross inspection, other pathologies may be indicated by aberrant weight, such as myocardial hypertrophy [27] and renal toxicity [28]. With the ability of spectral CT to non-invasively measure whole-organ mass through spectral physical density quantifications, mass measurements can expand beyond traditional post-mortem analysis to opportunistic screening of possible pathology in living patients. Given the accuracy and stability with different acquisition parameters, evaluation of mass may be applied to not only general opportunistic screening on existing scan protocols, but also characterization of solitary lesions. It covers a different marker for pathology, thus unlocking new diagnostic abilities.

Temperature evaluations with spectral physical density quantifications facilitate non-invasive, real-time temperature monitoring that can serve as real-time feedback to interventional radiologists. With ablation, tumors must reach a lethal threshold of 60 °C to be considered successful while ideally maintaining a safety margin from critical structures and sparing as much healthy tissue as possible [22]. Even with developments in ablation technologies, recurrence rates, such as with hepatocellular carcinoma [29]–[31], have remained higher than desired. As such, the addition of temperature monitoring presents an opportunity to reduce the recurrence rate by directly providing feedback and ensuring a complete ablation. Previous studies of CT thermometry have correlated HU to temperature linearly and quadratically with sensitivities ranging from -2.0 to -0.23 HU/°C in *ex vivo* tissues [32]–[35]. Furthermore, *in vivo* tissues have been utilized to investigate temperature distributions using the relationship of HU to temperature [36], [37]. However, HU incorporates both the effects of temperature and compositional changes while physical density isolates the effect of temperature changes without any approximations. By applying thermal volumetric expansion to spectral physical density quantifications, temperature can be estimated to determine whether the lethal threshold was reached for thermal ablation, thereby reducing incomplete ablations and local recurrences.

This study, however, had some limitations. First, validation of the physical density model illustrated differences in physical density not within manufacturer reported tolerance for the two lung inserts (LN 300, LN 450). This deviation in accuracy is likely due to non-uniformity of the insert rather than inaccuracies in the model [2]. Such uniformities resulted in large deviation in dual energy CT measurements, and thus the values were not within tolerance. Second, mass measurements were only performed on soft tissue and did not include tissues of higher density, i.e., bone. Moreover, the soft tissue utilized in this experiment, muscle, was generally more uniform compared to other soft tissues. Future experiments on a larger number of samples with tissue variability are planned for a follow-up study. Third, a lethal threshold of 60 °C was not reached in temperature experiments. Instead, temperature between 22 and 45.5 °C were achieved, which matched the required change in temperature to reach the lethal threshold from a normal body temperature of 60 °C. The change in temperature mimics what is expected in the *in vivo* scenario, suggesting its relevance and applicability. Fourth, *ex vivo* bovine muscle was utilized for temperature evaluations and does not reflect the same properties as liver, an organ that may contain lesions treated with ablation. Moreover, *in vivo* tissue differs from *ex vivo* tissue, such as the inclusion of the heat-sink effect and presence of perfusion [21], [22], [38]. Future studies will necessitate more precise evaluations in order to translate non-invasive mass and temperature evaluation applications to the clinic.

## V. Conclusion

Accurate and stable spectral physical density quantifications enable non-invasive mass and temperature measurement that can assist in diagnostic imaging for pathological detection and interventional radiology for thermal therapy monitoring. In particular, real-time non-invasive temperature measurements open an avenue for intraprocedural monitoring that will ultimately improve clinical care by providing feedback to interventional radiologists, thus decreasing the recurrence rate and ensuring minimal damage to critical structures for thermal ablation.

## Data Availability

All data produced in the present study are available upon reasonable request to the authors.

## Acknowledgment

The authors would like to thank Michael Geagan for his help with 3D-printing the guides.

